# Enhancing Translational Stroke Rehabilitation: Task-Specific Action Observation Therapy for Motor Network Optimization

**DOI:** 10.1101/2025.03.03.25323286

**Authors:** Paola Romano, Sanaz Pournajaf, Leonardo Buscarini, Elena Sofia Cocco, Peppino Tropea, Massimo Corbo, Marco Franceschini, Francesco Infarinato

## Abstract

**Background:** Action Observation Therapy (AOT) leverages mirror neuron system (MNS) activation to enhance neuroplasticity and motor recovery after stroke. While AOT has demonstrated positive effects, the neural mechanisms underlying its efficacy, particularly regarding task type and motor network engagement, remain unclear. This observational cohort study investigates cortical activation during the observation of goal-oriented Activities of Daily Living (ADLs) in healthy individuals and chronic stroke patients.

**Methods:** Twenty stroke survivors with right hemiparesis (≥6 months post-stroke) and 23 age-matched healthy controls observed standardized videos of finalized actions (FA), non-finalized actions (NFA), and control videos (CV). A time-frequency electroencephalography (EEG) signal analysis examined sensorimotor rhythm modulation during action observation (AO), Event-Related Spectral Perturbation (ERSP) analysis was used to quantify mu rhythm desynchronization (8-13 Hz) and beta suppression (14-25 Hz).

**Results:** Healthy individuals exhibited significant mu rhythm desynchronization, predominantly in the beta band, with stronger and prolonged responses to goal-directed actions (self-care and feeding). Stroke patients showed delayed and attenuated beta suppression, particularly in the affected hemisphere, but retained selective responsiveness to goal-oriented tasks. Rebound effects occurred in all categories in both groups but were most pronounced for NFA and CV, particularly in the beta band (e.g., *t*(44) = −4.28, *p* < 0.0001 and *t*(44) = −2.163, *p* = 0.04 in healthy individuals).

**Conclusions:** This study underscores the importance of task specificity in AOT, demonstrating that goal-directed actions optimize motor network engagement. Attenuated but preserved beta suppression in stroke survivors supports the integration of standardized ADLs into AOT protocols to enhance neuroplasticity and motor recovery. Findings highlight the potential for EEG-based biomarkers to monitor AOT efficacy and personalize stroke rehabilitation, with possible implications for digital health and telerehabilitation applications.

**Clinical Trial Registration:** Clinical Trial Registration - URL: http://www.clinicaltrials.gov. Unique identifier: NCT04047134.

## Introduction

The mirror neuron system (MSN) may be interpreted as an intrinsic mechanism that is not isolated but rather inherent to numerous motor-related cortical areas since many of the regions encoding a movement also respond to its observation^1–3^. Mirror neuron system-based training has become a prominent treatment technique in recent years, introducing a novel strategy for functional rehabilitation after stroke^4^. Upper limb recovery after stroke is crucial for increasing functional performance and participation in daily living^5^: effective strategies typically involve early, intensive, repetitive, and task-oriented practice^6^.

Recently, innovative strategies for rehabilitation have been proposed as an alternative to conventional physical therapy^7,8^. Among them, Action Observation Therapy (AOT) represents an innovative treatment that generates multisensory stimulation and promotes neuroplasticity through the activation of the MNS^9,10^. AOT consists of observing an individual performing a motor task, via video or in real life. For example, the stroke survivor is instructed to watch a video showing an adult stretching out his hand to pick up a cup, bringing it to his mouth, and then returning it to its initial position. After observing a repeated video sequence, the individual is asked to perform the same action^11^.

Findings from electroencephalogram (EEG) acquisition indicate a significant increase in event-related desynchronization power under action observation (AO) even after a single session in chronic stroke patients both with and without higher-brain dysfunction. These findings support the favorable outcomes reported with repetitive AOT in terms of motor rehabilitation and cortical plasticity^12^.

The analysis of the literature highlights heterogeneity in treatment protocols, such as video content, observation and execution times, perspective used, and treatment dosage^10,11^. Despite growing evidence, further research is needed on neural network reorganization, triggered by MNS activation after injury. To the best of our knowledge, current literature lacks studies comparing brain activity changes in motor areas and EEG oscillatory patterns during the observation of daily life gestures involving different household objects^13–15^. Research on cortical responses in healthy individuals and stroke patients while observing upper limb movements in videos is limited and primarily focuses on abstract object use. What is clearer is: (i) observing goal-directed actions is more efficient than non-goal-directed actions^16^; (ii) it is necessary to split the gesture into the simplest congruent movements to facilitate motor learning^17^; (iii) interaction with real household objects shows no differences between real demonstration and video settings^18^.

AO induced by manipulation of specific target objects (i.e., blue ball, stick, or cube) leads to suppression of sensorimotor mu rhythm in alpha and beta bands^18,19^. In particular, Angelini et al. (2018) and He et al. (2020) suggested greater beta motor rhythm recruitment in face-to-face interactions during AO in healthy subjects^19,20^. Studies on stroke patients, such as Zhu et al. (2019), highlighted greater activation in the primary motor cortex when observing and executing grasping movements, stressing the necessity of identifying the most appropriate strategies to enhance stroke patients’ performance^21^.

These studies suggest that future research should determine the optimal intensity, action categories, and treatment duration to ensure that improvements from action observation persist and generalize to other functional domains. So far, research has investigated the influence on cortical motor activity of object-directed actions subdivided into categorized Activities of Daily Living (ADLs) in individuals with stroke ^14^. However, further studies are needed to integrate current findings with additional research on object-oriented tasks in daily living contexts to enhance their effectiveness.

This observational cohort study aims to address these gaps in the current literature by investigating cortical activation elicited by well-selected, object-directed, and goal-oriented actions categorized as ADLs in healthy individuals and chronic stroke patients. Using time-frequency analysis of EEG signals, we seek to elucidate the mechanisms underlying neural activity modulation to inform the design of targeted therapeutic protocols that enhance functional recovery after stroke. The goal is to identify optimal strategies for stimulating neuroplasticity and promoting motor recovery through AOT, with potential applications in clinical practice and digital telerehabilitation platforms.

## Materials and Methods

This observational cohort study was carried out during an AO protocol administration displayed on video. Simple, short, and standardized actions from four different categories of ADLs in a neutral and uniform background were shown by employing household objects.

Able-bodied right-handed subjects over 45 years old without upper limb pathologies (peripheral neurological damage, inflammatory degenerative joint diseases, fracture, or trauma results), cognitive and/or severe visual deficit were included in the control healthy group (CG).

Patients in the experimental group (EG) were evaluated by Physical Therapists, and those with severe impairments were excluded. Eligibility criteria included adults affected by right hemiparesis after stroke in chronic status (onset ≥ 6 months), first unilateral ischemic stroke confirmed by Magnetic Resonance Imaging, segmental Modified Ashworth Scale -Upper Limb ≤ 2, moderate to mild upper limb motor impairment (Fugl-Meyer Assessment -Upper Limb > 22), Chedoke-McMaster Stroke Assessment > 1, and the ability to comprehend instructions and provide informed consent. Exclusion criteria included hemorrhagic stroke, bilateral dysfunction, severe sensory deficits in the paretic upper limb, severe cognitive impairment, aphasia, and apraxia.

The same AO protocol, recording systems (sampling frequency of 1000Hz using the EGI Geodesic 400 128-channel EEG and the BrainAmp 64-channel EEG systems), and EEG signals pre-processing described in the Franceschini et al. (2022) study^14^ were used to compare the cerebral activity modulation between healthy chronic stroke participants. During the single-day EEG recording protocol, subjects were asked to carefully observe actor gestures in an allocentric perspective^20,22^ on the 23-inch laptop monitor placed at about 50 cm of visual distance. The sequence of 40 video stimuli lasting around 4 seconds repeated three times underpin the experimentation, of which 32 human motion videos, showing an actor performing an upper limb motor task, and 8 Control Videos (CV), showing landscapes with the absence of persons^23^. The visualization presented to subjects was designed to maximize the effect of AO through a thoughtful choice of video stimulations. Human motion videos were divided into the following sub-categories: 24 Finalized Actions (FA) and 8 Non-Finalized Actions (NFA). FA includes manipulation with real objects and was further subdivided into:

- FEedings Actions (FEA), The actor executes nutrition-related tasks grasping edible foods;
- External Actions (EA), the actor performs target-oriented actions in the peripersonal space using objects;
- Self-Care Actions (SCA), the actor interacts with an object aimed at personal care (e.g., combing the hair).

In the FA, the actor (male or female) divided the movement into a first brief time interval to reach the object and a second one to finalize the action using it.

NFA contains gestures not finalized to a specific action (i.e. arm slow movements with fixed direction and without interaction with objects).

During the AO protocol, a black screen (evt1) as intertrial of duration was randomly extracted from a uniform distribution of values between 3 and 4, and a subsequent fixation cross of 500 ms duration (evt2) alternated with video stimuli (evt3).

The affected and unaffected motor cortices will be compared with the dominant and non-dominant hemispheres of healthy individuals, respectively. To ensure homogeneity and minimize bias, only right-handed healthy participants and individuals with right-sided hemiparesis were recruited. All subjects observed left-hand actions performed by the actor from a frontal perspective. This setup ensures that the left hemisphere (C3) corresponds to the dominant and affected Primary Motor Cortex (PMC), while the right hemisphere (C4) corresponds to the non-dominant and unaffected PMC in both healthy and stroke participants, respectively.

Event-Related Spectral Perturbation (ERSP) calculation was employed to determine cerebral rhythms modulation through the Morlet transformation: it returned the time-frequency map of event-related signal power relative to a baseline. The classical method to depict the rhythms of Event-Related Desynchronization (ERD)/Event-Related Synchronization (ERS) over time^24^ is overcome with time-frequency transforms for each accepted and cleaned epoch computed for electrodes in the frequency range of interest^19,25^. The result is a colored panel from which the behavior of the analyzed biomarker specific in time and frequency bins can be discussed. In the case of cerebral rhythms modulation, warm colors (red) indicate an increase in spectral power, and cold colors (blue) a decrease in it with respect to the baseline power.

Before spectrogram implementation, pre-processing and trial-by-trial segmentation were drawn on the methods described in Franceschini et al. (2022)^14^. Then, the analysis addressed the time-frequency analysis consisting of the ERSP map, power ratio, time course, and ERD/ERS. Each of them was referred to the primary motor area, identified by electrodes C3 and C4. Literature reported several time-frequency analysis methods^26^. In this study, the ERSP computation followed steps based on an in-house routine constructed on the instructions of Avanzini et al. study (2018)^19^. The number of cycles considered to calculate Morlet function w increases linearly with frequency in the range from 8 Hz to 25 Hz, with 3 cycles associated with the lowest frequency, up to 15.8 at the highest frequency. All wavelet-modulated and normalized trials were averaged to derive the signal power in the time-frequency domain for each video category. The relative event-related power trend (P_task_) was calculated as the ratio between the spectrogram in the evt3 time interval and the mean power (P_baseline_) along a 1500ms baseline positioned in the pre-stimulus period (in evt1). The power ratio is then calculated as follows:

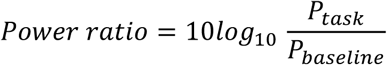

The reference level of the logarithmic scale is equal to 0, corresponding to the green value, which indicates no change in power between the interval under exam and the reference baseline. Expressing the map in these terms allows for immediate visual discrimination of what is happening to neuronal populations along with the action observation from the video beginning to the end.

Finally, analysis focused on the time-frequency indices calculation to derive the temporal modulation of the α (range 8-13 Hz) and β (range 14-25 Hz) cerebral rhythms^27^: α and β bands are associated with sensorimotor processing and action observation, the main cortical functions elicited by the MNS activation, and contain the two dominant frequencies of EEG µ rhythm, investigated in the literature studies when searching a neurophysiological marker of mirror neuron activity, reflecting action understanding, and motor resonanceof observed actions performed by others^15,28,29^. Finally, to extract time-frequency indices and assess desynchronization in the α and β bands, the averaged spectrogram was first calculated for each subject and video category, focusing on the left and right sensorimotor cortical areas (respectively C3 and C4 signals). From this, an averaged frequency curve was generated, and the two minima were identified, representing the maximum desynchronization in the α and β ranges. For each subject, the frequency bins and bands (3 Hz wide) of interest at these desynchronization peaks were selected, and the spectrogram was averaged within these intervals resulting in time desynchronization curves for the α and β bands were then divided into eight intervals spanning the range [0; 4] s.

Two graphs were produced for each cortical area and subject—one for the α rhythm and one for the β rhythm—each displaying eight points corresponding to the average power in the selected time intervals. The studied indices, labeled from T1 to T8, along with the maximum desynchronization values and their corresponding frequencies, were extracted for the entire video duration. A comparison was conducted within the CG and EG separately, following the same analysis algorithm. However, for consistency in comparing between the two groups, the frequency ranges for the α and β bands were predefined as 8-13 Hz and 14-25 Hz, respectively.

### Statistical analysis

Both the Kolmogorov-Smirnov and Shapiro-Wilk tests confirmed the normality of the data across all case combinations. A cross-validation was performed using non-parametric tests for the few combinations where one of the five categories of the independent factor displayed a slight deviation from normality. Levene’s test indicated no violation of the assumption of homogeneity of variances. Consequently, an ANOVA was employed with within-subjects factors set to channel (C3 and C4), band (alpha and beta), index (T1 to T8), and the independent variable set to video category, to analyze the data for healthy and pathological subjects separately. Additionally, a t-test was conducted under the same case subdivisions, with Group (CG and EG) as the independent variable, to perform between-subjects comparisons.

### Data Availability

The data supporting the findings of this study are available from the corresponding author on request, and will be available in Zenodo Open Research Repository.

## Results

### Participants

Twenty-three right-handed healthy participants (mean age ± SD = 59.8 ± 10.9 years) without upper limb pathologies (e.g., peripheral neurological damage, inflammatory degenerative joint diseases, fractures, or trauma outcomes), cognitive impairments, or severe visual deficits were included in the control group (CG). Twenty stroke survivors with right hemiparesis in the chronic phase (stroke onset ≥ 6 months; mean age ± SD = 71.7 ± 10.8 years) who met the inclusion criteria were recruited for the experimental group (EG). The study flow chart, available in the supplemental material, illustrates the participant selection process, exclusions, and data acquisition details. A demographic and clinical description of the stroke sample is reported in Table 1.

**Table 1:** Demographic and clinical description of stroke survivors with right hemiparesis subjects (N=20)

|  |  |
| --- | --- |
| **Age (years)** | 71.7±10.8 |
| **Gender (female/male) %** | 8 (40%) /12 (60%) |
| **MAS-UL** | 1.25 (0-6) |
| **FMA-UL** | 51 (29-66) |
| **FAT** | 5 (1-5) |
| **mBI** | 94 (46-100) |

*Mean ± standard deviation; N (%); median (minimum–maximum). Modified Ashworth Scale-Upper Limb (MAS-UL); Fugl-Meyer Assessment-Upper Limb (FMA-UL); Frenchay Arm Test (FAT); and modified Barthel Index (mBI).*

### Cortical Rhythm Reactivity During AO

The results are presented separately for the alpha and beat bands, which represent the two subcomponents of mu rhythm suppressed during MNS activation. Figures 1 and 2 show ERSP maps for both groups, revealing distinct patterns of sensorimotor rhythm modulation during AO. All FA categories elicited an evident central visuomotor reactivity compared to NFA, and especially CV in both EG and CG.

**Figure 1:**
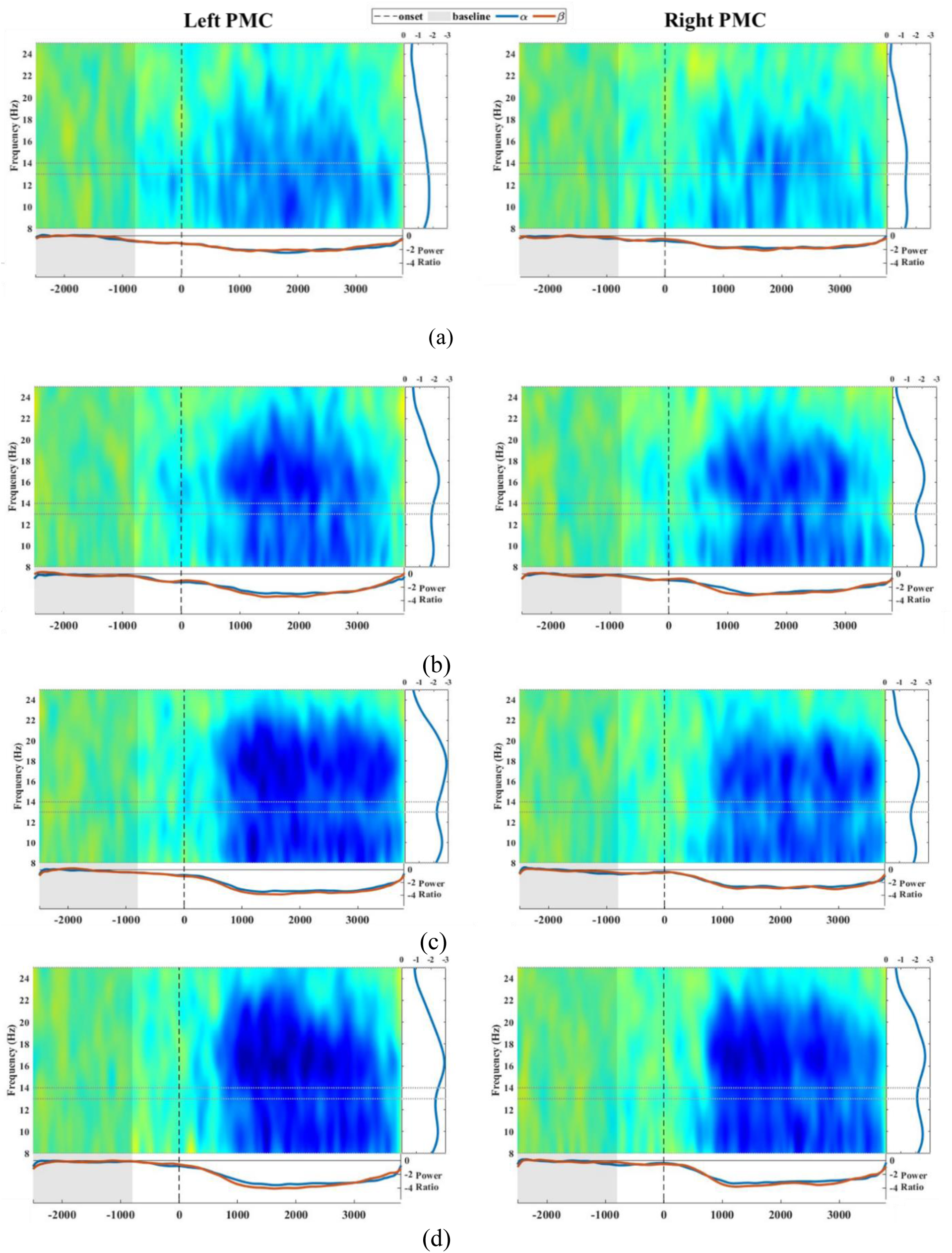
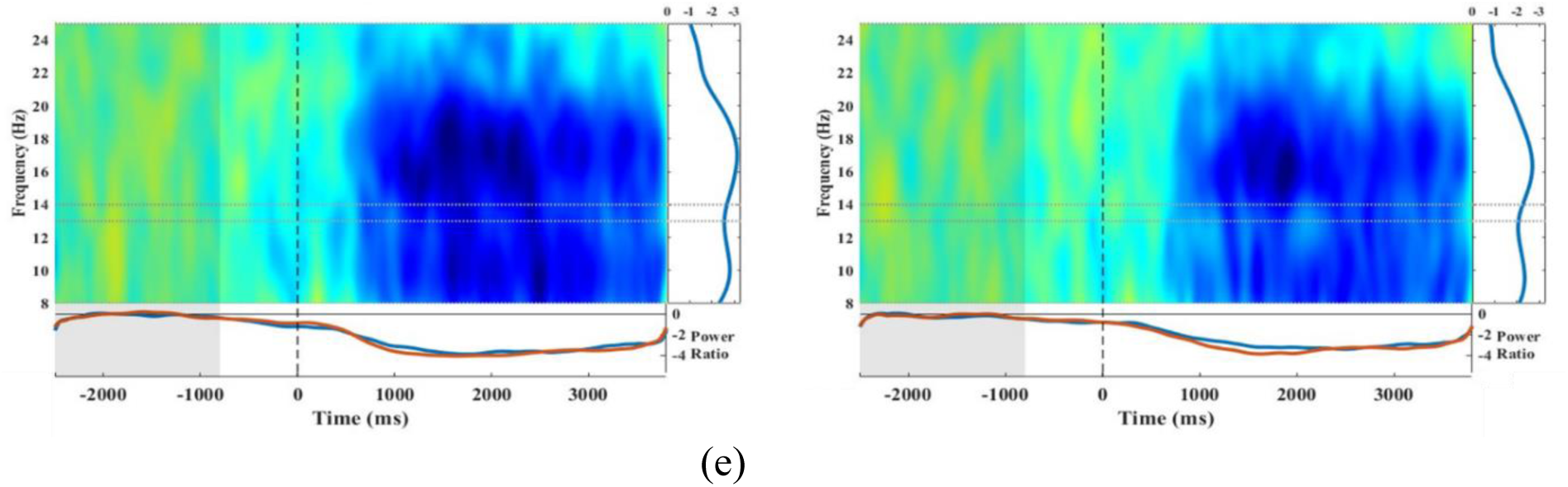
ERSP from dominant (left) and non-dominant (right) PMC of examined healthy sample during action observation averaged across (a) CV, (b) NFA, (c) FEA, (d) EA, (e) SCA trials. The X-axis represents time in ms starting from the lower extremity of baseline (−2300 ms) to the fixed end of the trial (4000 ms). The central panel is the depiction of ERSP calculated by the Wavelet algorithm. The dotted line represents the onset of stimulus evt3 (0 s). The vertical right plot represents the power ratio averaged over evt3. The horizontal plots show the average over the significant frequency bins of α (blue) and β (red) rhythms.

**Figure 2:**
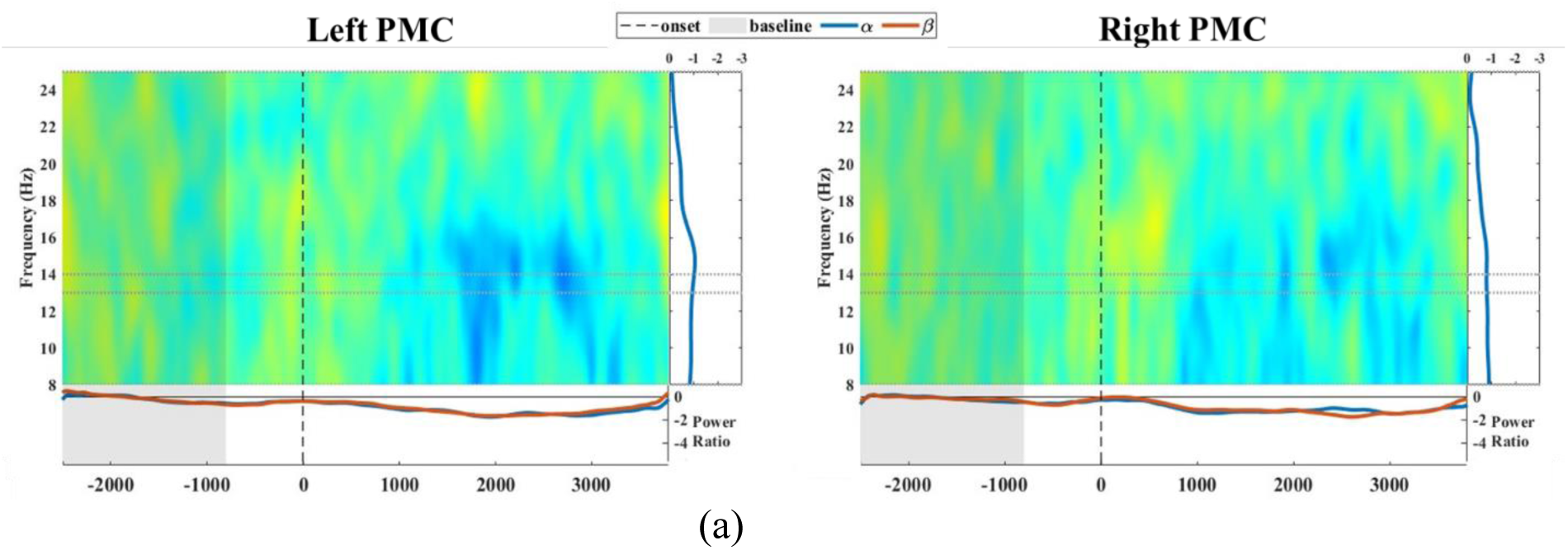
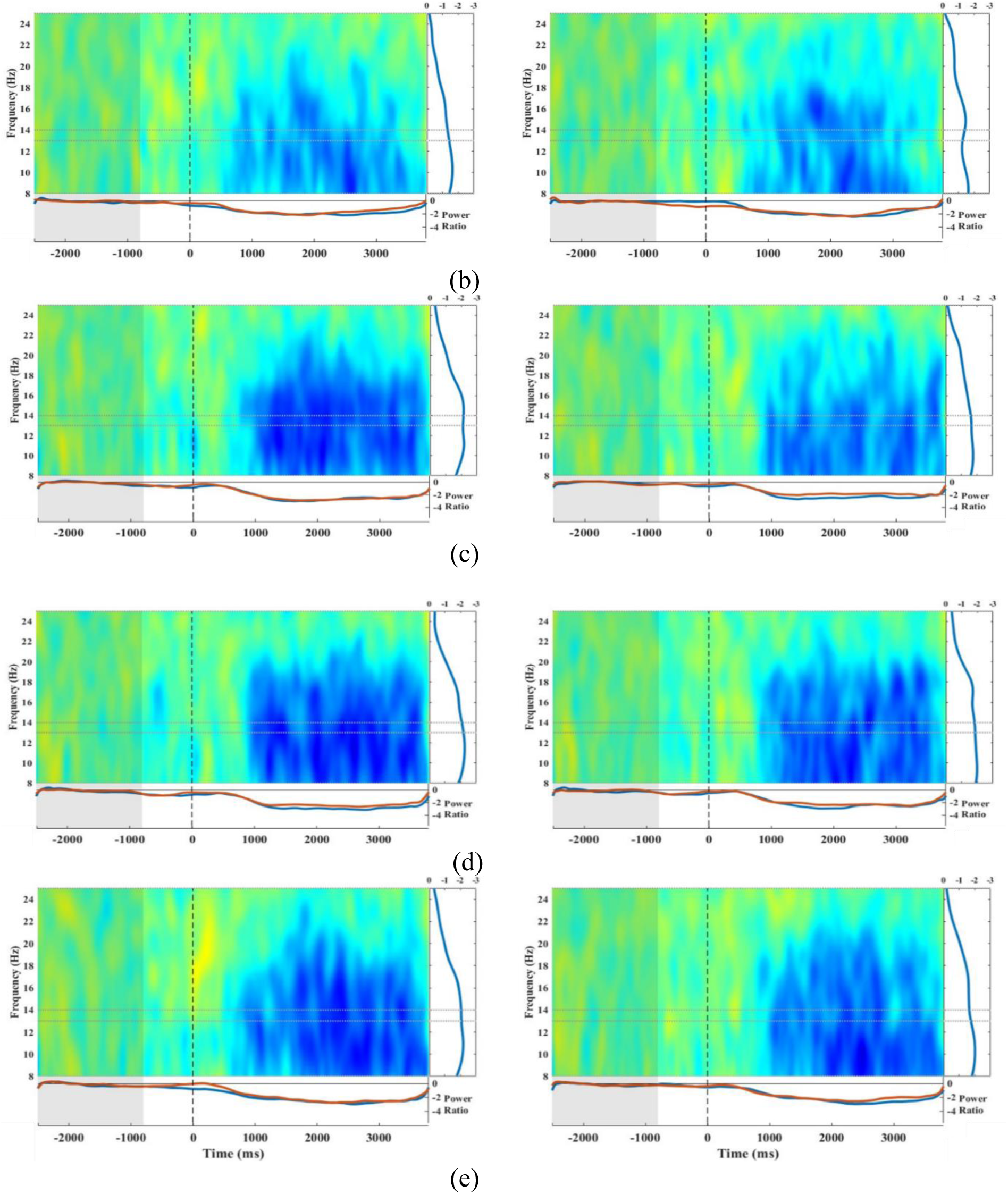
ERSP from left (affected) and right (unaffected) PMC of examined stroke participants during action observation averaged across (a) CV, (b) NFA, (c) FEA, (d) EA, (e) SCA trials. The X-axis represents time in ms starting from the lower extremity of baseline (−2300 ms) to the fixed end of the trial (4000 ms). The central panel is the depiction of ERSP calculated by the Wavelet algorithm. The dotted line represents the onset of stimulus evt3 (0 s). The vertical right plot represents the power ratio averaged over evt3. The horizontal plots show the average over the significant frequency bins of α (blue) and β (red) rhythms.

#### Healthy Participant

Healthy people exhibit more evident central visuomotor reactivity; they suppress EEG rhythms more rapidly after evt3 onset. Furthermore, β desynchronization appears distributed across both low and high beta bands, and action observation induces a less pronounced desynchronization in the α frequency range.

#### Stroke Participants

β desynchronization was weaker and delayed, predominantly limited to lower β frequencies, while α band desynchronization was more pronounced. This reflects altered motor network reactivity post-stroke.

### ANOVA and Pairwise Comparisons

Significant differences in ERSP were identified through ANOVA with Bonferroni post hoc tests, as summarized in Tables 2 and 3 for each sub-interval in investigated frequency bands and channels. From T_3_ to T_8_, both groups show significant differences in pairwise comparisons. Specifically, the key findings include:

1. Group-Specific Patterns:

- In healthy participants (CG), β desynchronization was stronger and more consistent in the dominant PMC across FA categories, particularly for Feeding Actions (FEA) and Self-Care Actions (SCA) from T_3_ to T_8_ included.
- In stroke participants (EG), α band effects dominated, with significant beta suppression evident only during the later reaching and completing action phases (T_4_/T_5_ and T_8_).
2. Group-Shared Patterns:

- FA vs. CV and NFA: Both groups demonstrated significant differences between FA categories and CV, as well as between FA and NFA at T8 in the β band. FA consistently influenced dominant motor area activation until the end of the video.
- Bi-lateralization: Hemispheric differences during action observation emerge only under specific conditions in both groups, particularly for the FEA category at T4 in the β band (t = −2.140, df = 44, p = 0.038) for CG and T5 in the β band (t = −2.062, df = 38, p = 0.046) for EG, highlighting a noteworthy bilateral activation of the mirror neuron system.

**Table 2:**
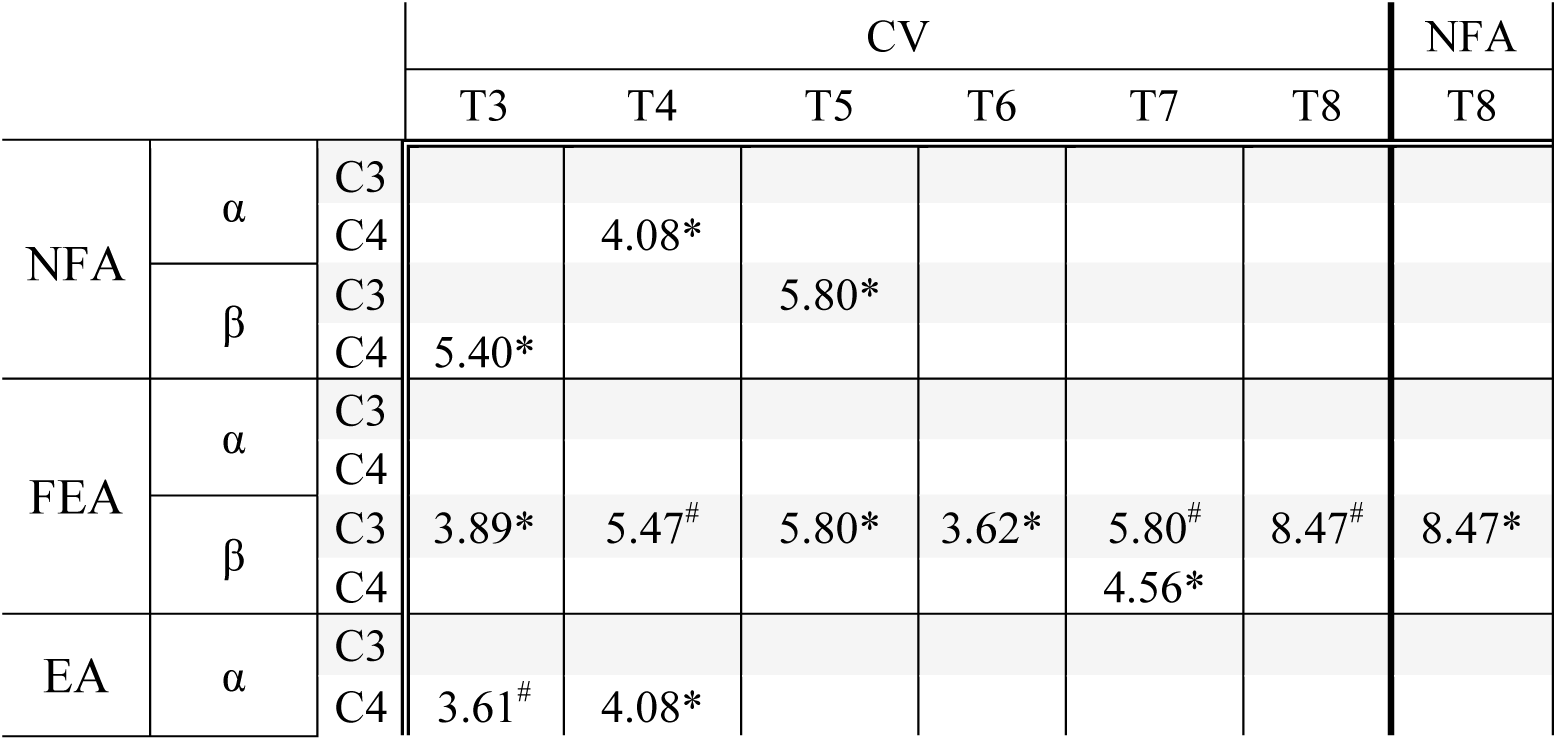
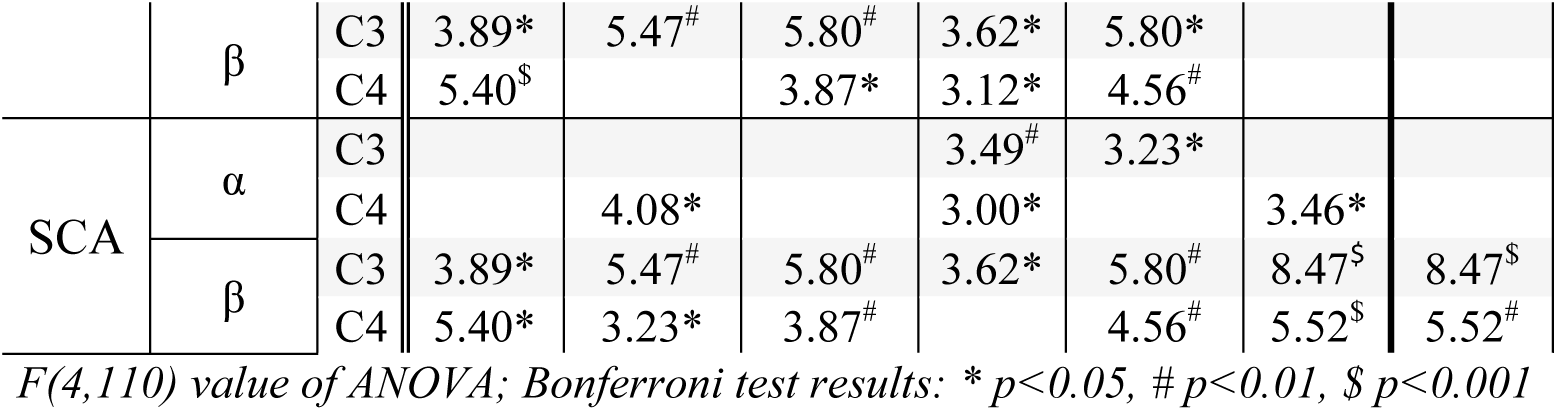
Pairwise comparisons between videos categories from ANOVA and Bonferroni post-hoc test of the time-frequency 8 indices during action observation in CG.

**Table 3:** Pairwise comparisons between video categories from ANOVA and Bonferroni post-hoc test of the time-frequency 8 indices during action observation in EG.

|  |  |  | CV |  |  |  |  |  | NFA |
| --- | --- | --- | --- | --- | --- | --- | --- | --- | --- |
|  |  |  | T3 | T4 | T5 | T6 | T7 | T8 | T8 |
| NFA | $\alpha$ | C3 | | | | | | | |
|  |  | C4 |  |  |  |  |  |  |  |
| | $\beta$ | C3 | | | | | | | |
|  |  | C4 |  |  |  |  |  |  |  |
| FEA | $\alpha$ | C3 | 3.39* | 3.78<sup>#</sup> | | | | 5.51* | |
|  |  | C4 |  |  |  |  |  |  |  |
| | $\beta$ | C3 | | 2.53* | | | | 6.25* | 6.25* |
|  |  | C4 |  |  |  |  |  |  |  |
| EA | $\alpha$ | C3 | 3.39* | 3.78* | | 3.39* | | 5.51<sup>#</sup> | |
|  |  | C4 |  | 2.65* | 3.72<sup>#</sup> |  |  | 4.99* |  |
| | $\beta$ | C3 | | | | | | 6.25* | 6.25* |
|  |  | C4 |  |  | 3.20* |  |  |  |  |
| SCA | $\alpha$ | C3 | | | | 3.39* | | 5.51* | |
|  |  | C4 |  |  | 3.72* | 4.16<sup>#</sup> |  | 4.99* |  |
| | $\beta$ | C3 | | | | | | 6.25<sup>#</sup> | 6.25* |
|  |  | C4 |  |  | 3.20* |  |  |  |  |

*F(4,95) value of ANOVA; Bonferroni test results: \* $p < 0.05$ , # $p < 0.01$*

Thus, we can affirm that the FA category consistently influences the motor areas activation until the end of the video visualization.

### Rebound Effects and Beta Desynchronization Loss

Figure 3 illustrates four panels depicting the different rhythmic modulation of the eight time-frequency indices from CG (first column) and EG (second column). Bonferroni post-hoc results are detailed in Table 4, which accentuates the most substantial loss of motor area stimulation in stroke participants in the beta band and the lesioned hemisphere. Beta suppression in stroke participants is significantly reduced across the entire video categories duration.

**Figure 3:**
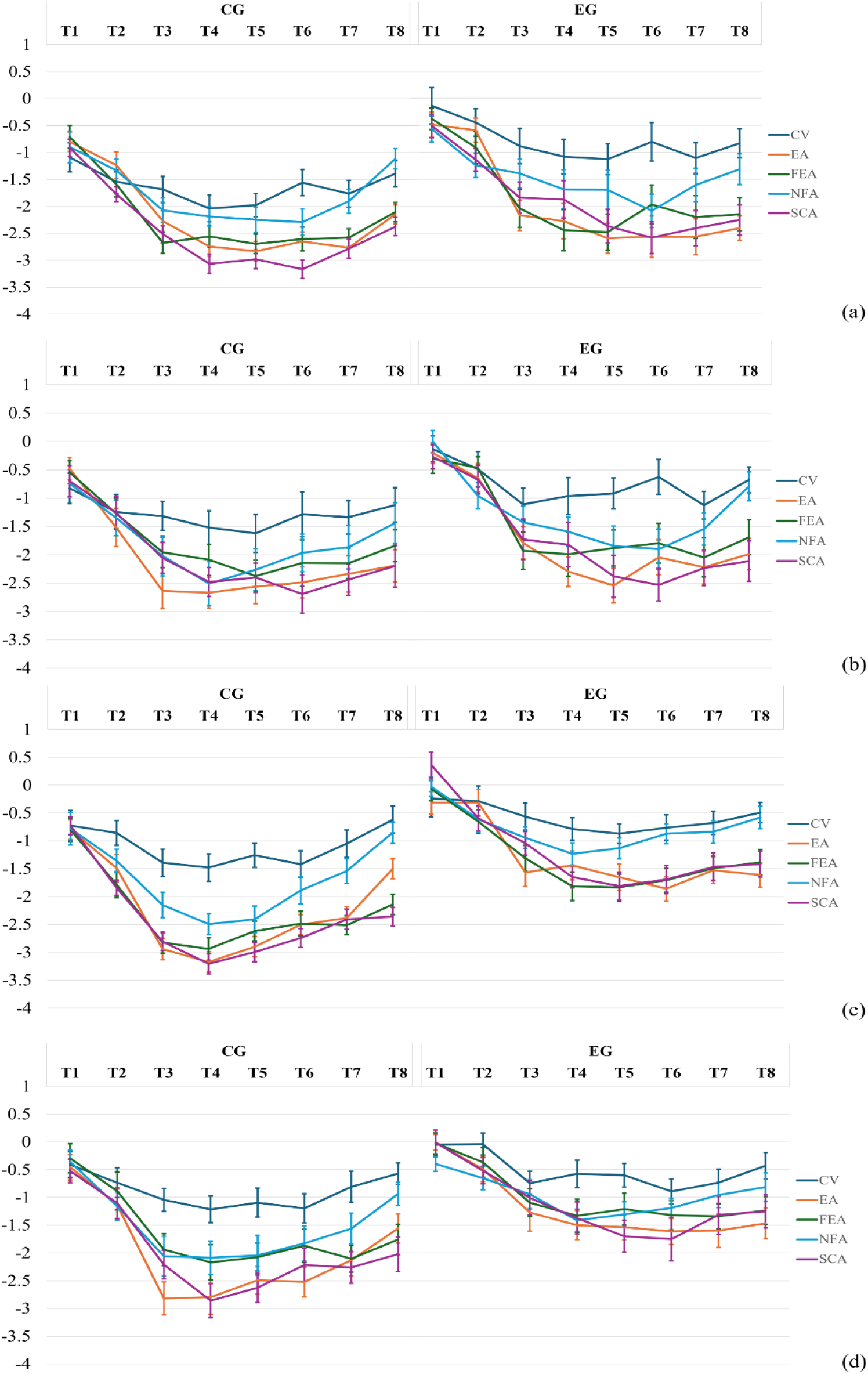
Trends (mean ± standard error) of time-frequency 8 indices in α band from C3 (a) and C4 (b), and in β band from C3 (c) and C4 (d) during action observation for healthy (first column) and stroke (second column) subjects.

**Table 4:**
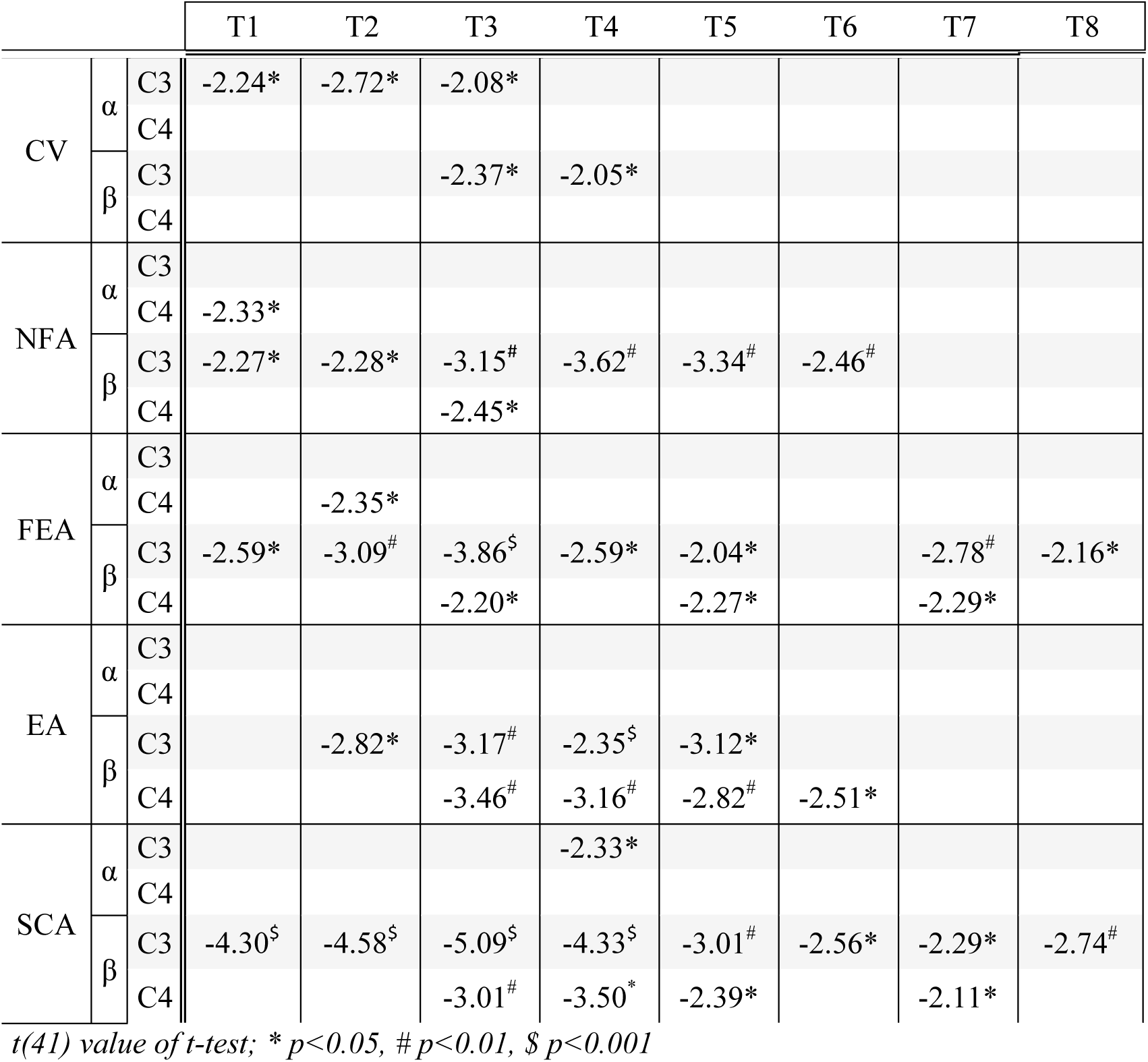
Significant pairwise comparisons between the two groups from the t-test under all conditions of cerebral rhythm, channel, and video category.

Beta suppression visibly diminished towards the end of videos in both groups, with notable rebound effects^21^ in the NFA category:

- In healthy participants (CG), NFA elicited a significant loss of rhythm suppression between T_4_ and T_8_ in the beta band (dominant: t(44) = −4.28, p < 0.0001; non-dominant: t(44) = −2.98, p = 0.005). Alpha band rebound effects were also significant (dominant: t(44) = −2.45, p = 0.018; non-dominant: t(44) = −2.08, p = 0.043).
- In stroke participants (EG), rebound effects were less pronounced but significant in the alpha band for the unaffected hemisphere (t(38) = −2.46, p = 0.018) and in the beta band for the affected hemisphere (t(38) = −2.47, p = 0.018).

Other video categories in CG also show significant effects in the β band between T_4_ and T_8_, including CV from the non-dominant, t(44) = −2.163, p = 0.04, EA from the dominant, t(44) = −4.03, p < 0.0001, and the non-dominant, t(44) = −2.74, p = 0.009, FEA from the dominant, t(44) = −2.37, p = 0.022 hemispheres.

Notably, no rebound was observed in SCA, and beta desynchronization during T8 remained significantly stronger for SCA compared to EA in CG (ANOVA: F(2,66) = 3.58, p = 0.028), performing test on data filtered for goal-directed actions only.

### Beta Peak Frequency Shifts in Stroke Participants

Stroke patients exhibited a significant shift in beta desynchronization peak frequency compared to healthy participants: EA at C3, 16.6 Hz in CG vs. 15.4 Hz in EG; FEA at C4, 17.2 Hz in CG vs. 15.8 Hz in EG, with borderline significance (t(41) = 2.01, p = 0.053); FEA at C3, a significant shift (17.5 Hz in CG vs. 15.3 Hz in EG) was observed (t(41) = 3.77, p = 0.001).

## Discussion

This observational cohort study investigated the reactivity of the sensorimotor rhythms - specifically, the alpha and beta components of the mu rhythm, which are associated with both action perception and execution-related neural processes - over the central scalp during the observation of video stimuli depicting different action categories in both healthy individuals and stroke patients. By examining the neurophysiological responses, this work offers valuable insights that can be applied to enhance clinical practice. Notably, to the best of our knowledge, no previous EEG studies have specifically focused on functional ADL-related actions ^13–15^. This study addresses this gap in literature, contributing to a better understanding of sensorimotor dynamics in both typical and pathological populations. Mu rhythm desynchronization, widely regarded as an indicator of neural mirroring activity, reflects motor cortical engagement during AO^30^. However, the quantitative extent of this recruitment remains underexplored, particularly in stroke populations^31^. Visual stimuli based on ADLs have been effective in reorganizing damaged cortical areas, as demonstrated in previous studies where AOT using real-life gestures stimulated functional recovery in post-stroke individuals^11,32,33^.

To further understand this phenomenon, a time-frequency analysis of EEG signals examined motor activity modulation throughout 4-second videos of simple, short object-directed actions (FA) compared to neutral actions (NFA) and control stimuli (CV) depicting landscapes. Recorded videos of 4 seconds per action are consistent with protocols shown in prior studies to optimize AOT efficacy through repetition ^34^.

To partially avoid potential lateralization biases, left-hand actions were shown in a mirror-view perspective^34^, involving right-handed healthy participants and stroke patients with right hemiparesis, aligning with evidence that lateralization effects can influence MNS activation^35,36^. Moreover, the employment of the significant frequency bin, representing the frequency value with maximal desynchronization^37^, minimized inter-subject variability, thereby enhancing the accuracy of within-group analyses. These methodological approaches allowed for a more precise investigation of the temporal and spectral dynamics during AO, leading to findings that partially diverged from those observed in our prior research^14^.

Functional actions involving object manipulation were consistently more effective in activating motor cortical areas compared to neutral gestures without manipulation and control videos depicting nature, in both groups. Notably, self-care actions (SCA) showed the most prolonged and significant effects. Stroke participants exhibited delayed and weaker beta rhythm suppression, particularly in the lesioned hemisphere, but demonstrated preserved reactivity to goal-directed actions. These findings underscore the potential of incorporating standardized FAs with a neutral background into Action Observation Therapy protocols to optimize motor network engagement and promote functional recovery.

From time-frequency maps and the derived biomarkers in healthy people, results showed a significant mu rhythm reactivity starting around 1 second after the video began for all finalized action categories concerning the control videos. Meaningful effects emerge mostly in the β band, in the dominant motor cortex for feeding actions (FEA), and bilaterally for the remaining finalized actions. It is known from the literature that effective interaction with objects is not necessary to activate MNS in humans^1^, and also intransitive actions could induce motor resonance; anyway, when a gesture is merely suggested rather than goal-oriented, the resynchronization of neural rhythms occurs more rapidly after a more subtle neuronal activity. This is further supported by what occurs at the end of the video (T_8_), where both FEA and SCA sustain more intense neuronal recruitment compared to NFA, even in statistical terms. The stronger frontal alpha and beta desynchronization around the final reaching phase (T4-T6 sub-intervals) is noticeable from figures and statistic tests; whereas, rebound phenomena^38^, i.e. the loss of the rhythms suppression magnitude at the final part of the stimulus, appears during the completion phase, also for the goal-oriented tasks and mostly evident for actions involving the peripersonal space (EA): at T_8_ SCA returns significant differences compared to EA. With these considerations, we can conclude that FA and especially SCA sustain desynchronization for a longer duration, although it decreases.

Stroke-induced plasticity increases through ipsilateral pathways promoting compensatory activity in the unaffected hemisphere and supporting motor recovery and functional adaptation^36,37^; indeed, the most statistically significant post-hoc results in the pairwise comparison between actions and control videos are observed in this area. Event-related modulation of the neural processes in alfa and beta bands is a biomarker of chronic state status and recovery^41^. Motor areas in both the affected and unaffected hemispheres exhibited reduced rhythms suppression during AO. In the earlier sub-intervals of finalized actions observation, a pronounced alpha response was observed, followed only at the end of the videos by a significant beta suppression compared to the resynchronized rhythms under CV stimulation. This finding aligns with evidence from the literature showing that stroke patients exhibit a significant bilateral reduction in beta power, regardless of the lesioned hemisphere or lesion location^42,43^. Additionally, intransitive actions videos prompted a recovery in cerebral rhythm power at the end of the videos, consistent with results observed in the healthy control group. Statistical evaluation showed stronger mu suppression at C3 (in the contralateral hemisphere to the observed moving hand) than C4 during feeding actions, particularly midway through the video at the food grasping phase. Feeding videos featured repetitive, simple gestures, unlike those in the other two goal-directed action categories, likely driving contralateral hemisphere activation. Motor lateralization in unimanual tasks varies with complexity, with more complex actions involving bilateral hemisphere activation^44^. This could suggest unilateral upper-limb action observation induced contralateral μ desynchronization based on video category^45^. Anyway, in literature inconsistencies concerning mu lateralization during AO emerged^46^: the same experimentation selecting videos with the left moving hand could be necessary to corroborate this finding and avoid the hypothesis about the hemisphere dominance influence.

Between-group pairwise comparisons confirmed the loss of cortical networks after stroke event in chronic status, resulting in a decrease of activity magnitude from motor areas. From time-frequency panels, a loss of the high beta rhythm clearly appears for all categories: the extension and intensity of blue portions in healthy panels are with evidence reduced in stroke panels. The impact of video stimuli on MNS activation seems delayed for stroke patients observing colored maps. Statistical tests applied to all transitive and intransitive actions showed significantly notable results in the beta band for the dominant, and thus the lesioned, hemisphere. Furthermore, the weaker mu suppression in C3 in stroke participants is also validated from the statistically lowest frequency bin at which maximum desynchronization occurs both in the left and right hemispheres considering videos performing goal-directed actions. In line with the literature, in stroke patients, the most consistent changes have been reported in the beta frequency band in the lesioned hemisphere^42,43^. The goal of promoting brain reorganization to support functional recovery through neuroplasticity enhancement seems achievable by targeting beta rhythm, which is integral to mental activity and attention but is notably diminished in stroke patients. Finally, recorded stimuli videos with a face-to-face perspective promote higher recruitment of beta motor rhythm^20,47^.

Therefore, incorporating these video categories into an AOT protocol shows promise for clinical application by inducing beta suppression to enhance motor skill learning^47^. Despite modest activations, the results confirm the videos’ utility in rehabilitation and support the hypothesis of facilitating neuroplasticity. This study provides new tools to advance clinical practices by enhancing AOT’s benefits, which have been largely, although empirically, demonstrated in both acute and chronic stroke recovery.

## Conclusions

This study highlights the potential of goal-directed actions, particularly Self-Care Actions and Feeding Actions, in eliciting sustained motor cortical activation through action observation. These findings provide a strong rationale for integrating these tasks into action observation therapy protocols for post-stroke rehabilitation. By utilizing the robust beta rhythm suppression observed in healthy individuals and the delayed but responsive beta desynchronization in stroke survivors, AOT can be tailored to enhance neuroplasticity and motor recovery, even in chronic phases of stroke.

The study underscores the importance of task specificity in AOT design, demonstrating the limited efficacy of non-finalized actions in engaging motor cortical networks. Future protocols should prioritize ecologically valid, goal-directed tasks to maximize therapeutic outcomes. Additionally, the identification of alpha and beta rhythms as biomarkers of motor network engagement opens avenues for real-time monitoring and personalization of therapy using EEG-based tools.

Translationally, this research supports the adaptation of standardized AO stimuli into digital health platforms and telerehabilitation systems, enabling remote and accessible delivery of therapy. The structured and validated protocol proposed here addresses gaps in the variability of AOT designs, offering a foundation for future multicenter trials.

In conclusion, by targeting beta rhythm suppression and optimizing task-specific interventions, this study provides actionable insights for bridging the gap between neuroscience research and clinical practice, ultimately improving outcomes for individuals with stroke and other neurological conditions.

## Abbreviations (in alphabetic order)

ADLs: Activities of Daily Living
AO: Action Observation
AOT: Action Observation Therapy
CV: Control Videos
EA: External Actions
EEG: Electroencephalography
ERD: Event-Related Desynchronization
ERS: Event-Related Synchronization
ERSP: Event-Related Spectral Perturbation
FA: Finalized Actions
FEA: FEedings Actions
NFA: Non-Finalized Actions
MNS: Mirror Neuron System
PMC: Primary Motor Cortex
SCA: Self-Care Actions

